# Fertile-window misclassification in period-tracking applications and associated pregnancy risk: a large observational analysis

**DOI:** 10.64898/2026.02.12.26346180

**Authors:** Erica Brondolin, Bruno Hadengue, Danielle Perro, Kristina Gemzell Danielsson, Ingrid Granne, Brian T Nguyen, Dustin Costescu, Elina Berglund Scherwitzl, Raoul Scherwitzl, Kerry Krauss, Eleonora Benhar

## Abstract

**Objectives:** Given the widespread use of period-tracking applications and evidence that some users rely on fertile-window predictions for pregnancy prevention, we aimed to quantify pregnancy risk arising from misclassification of biologically fertile days by period-tracking applications, and to compare this risk across calendar-based and basal body temperature (BBT)–supported period tracking and a digital contraceptive regulated as a medical device.

**Methods:** We conducted an observational analysis of cycles of mobile fertility application users who logged urinary luteinizing hormone (LH) tests. Biologically fertile days were defined using an LH-based reference fertile window (days −5 to 0 relative to ovulation). Three approaches were evaluated: a calendar-based period tracking application, a BBT-supported period tracking application, and a FDA-cleared digital contraceptive. Outcomes included day-specific frequency of fertile days misclassified as safe, cycle-level misclassification, and predicted pregnancy risk per cycle. Analyses were repeated in a subgroup of irregular cycles.

**Results:** 543,167 menstrual cycles with a clear LH surge signature were included in the analysis. Calendar-based period tracking frequently misclassified fertile days as safe, with 67% of cycles containing at least one at-risk day and 25% containing at least one high-risk day. The mean predicted pregnancy risk per cycle was 22%, increasing to 65% in irregular cycles. BBT-supported period tracking reduced misclassification but remained associated with substantial risk (41% of cycles with at least one at-risk day; mean predicted pregnancy risk 9%). In contrast, the digital contraceptive showed consistently low misclassification (3% of cycles with any at-risk day and a mean predicted pregnancy risk of 0.5%).

**Conclusions:** Both calendar-based and BBT-supported period-tracking applications not intended for contraception frequently misclassify biologically fertile days and should not be considered reliable tools for pregnancy prevention. Regulated digital contraceptives demonstrate substantially lower pregnancy risk.

**Short condensation:** Period-tracking apps frequently misclassify fertile days as safe, including days with high pregnancy risk. In a large real-world analysis, both calendar- and BBT-supported trackers showed substantial risk, unlike digital contraception methods regulated as a medical device.

## Introduction

Period-tracking applications (sometimes also referred to as fertility-tracking applications) have become ubiquitous among women of reproductive age, with uptake particularly high among adolescents and young adults [1]. These tools offer convenient cycle monitoring and easily accessible information about menstruation and fertility, addressing long-standing gaps in reproductive health education. For many users, period tracking applications serve as an entry point into understanding their own reproductive physiology, and their widespread adoption reflects both growing interest in menstrual health and the increasing integration of digital tools into everyday self-care [2].

Despite these potential benefits, the rapid expansion of period-tracking technologies has raised concerns within clinical and public health communities. In particular, some applications present fertility-related predictions — such as estimated ovulation dates and fertile windows — that may function as de facto medical guidance, despite lacking validated algorithms capable of reliably identifying all fertile days. A recent global analysis found that menstrual tracking apps have been downloaded and used at large scale worldwide (more than 200 million downloads), and users engage with these tools for a variety of reasons beyond simple cycle logging, including about 8% using them to avoid getting pregnant [1]. Although most period-tracking apps are neither designed nor validated for contraceptive purposes, users may interpret features such as predicted ovulation dates or displayed fertile windows as reliable indicators of when conception is likely or unlikely to occur [3]. Figure 1 illustrates a generic example of how a typical period-tracking application presents predicted fertile days and ovulation timing to users.

**Figure 1.**
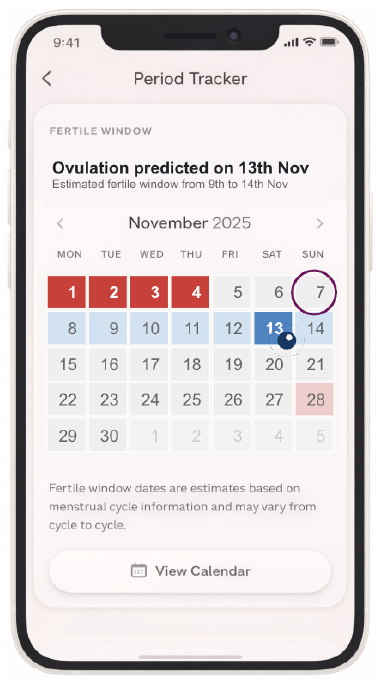
Illustrative example of a generic period-tracking application interface displaying menstruation days (red), predicted fertile window (light blue) and ovulation timing (dark blue). The purple circle indicates the current day. The figure is illustrative and does not correspond to any specific commercial product.

A 2025 study reported an increase in abortion-seeking individuals in the United Kingdom and noted a parallel rise in period-tracking app use among these patients [4]. Although this association does not establish causality, it raises the possibility that reliance on unvalidated fertile-window predictions may contribute to unintended pregnancies at the population level, warranting further investigation [5].

Quantitative evidence assessing the contraceptive risks of relying on period-tracking apps to guide intercourse timing remains limited. Most apps generate ovulation predictions primarily from calendar data, while some incorporate physiological biomarkers—most commonly basal body temperature—to refine these estimates. However, regardless of the underlying method, these tools typically present a narrow fertile window aimed at identifying the most fertile days, rather than all days on which conception is possible, and do not account for the well-documented variability in ovulation timing or cycle irregularity [6]. Because period-tracking apps are generally classified as wellness devices rather than medical devices, they are not required to undergo safety testing, algorithmic validation, or contraceptive effectiveness studies. Consequently, neither users nor clinicians can reliably assess how often predicted fertile windows fail to capture a woman’s true fertile period, potentially exposing individuals to unintended pregnancy risk when these predictions are relied upon without additional contraception.

Using a large real-world dataset of menstrual cycles, we estimate how often the fertile window predicted by typical period-tracking apps and by a digital contraceptive would fail to overlap with the biologically observed fertile window, identified using home urine luteinizing hormone tests (LH tests). Such mismatches represent periods in which a woman may believe she is not fertile when, in fact, she is—thereby creating a potential risk of unintended pregnancy if no additional contraception is used.

## Materials and methods

### Study design and data source

This study was an observational analysis of prospectively collected menstrual cycle data recorded via the digital contraceptive application Natural Cycles [7,8]. Natural Cycles is a regulated medical device, FDA-cleared in the United States and CE-marked in Europe for purposes of contraception.

The dataset consisted of anonymised daily records from users who had provided informed consent for the use of their data for scientific research. Users logged body temperature measurements (used as a proxy for basal body temperature) and menstruation dates and could optionally record results from home urine luteinizing hormone (LH) tests.

The earliest user registration date among included users was 1 September 2018, and the dataset was extracted on November 10th, 2025. No intervention, feedback, or contact with users occurred as part of this study.

We obtained an ethics waiver for the analysis of de-identified data (Reading Independent Ethics Committee, 10022023).

### Participants and cycle selection

From all cycles recorded in the application, we selected cycles with a total length of 15–90 days, excluding cycles occurring within the first three cycles after hormonal contraception or pregnancy. To define a reference dataset with well-defined LH surges, cycles were included only if a valid LH surge was identified, defined as positive LH tests recorded on one or two consecutive days with no other positive LH tests in the same cycle. The identified LH surge was verified by requiring the resulting luteal phase length to fall within ±2 days of the population average luteal phase length [9] (Figure 2).

**Figure 2.**
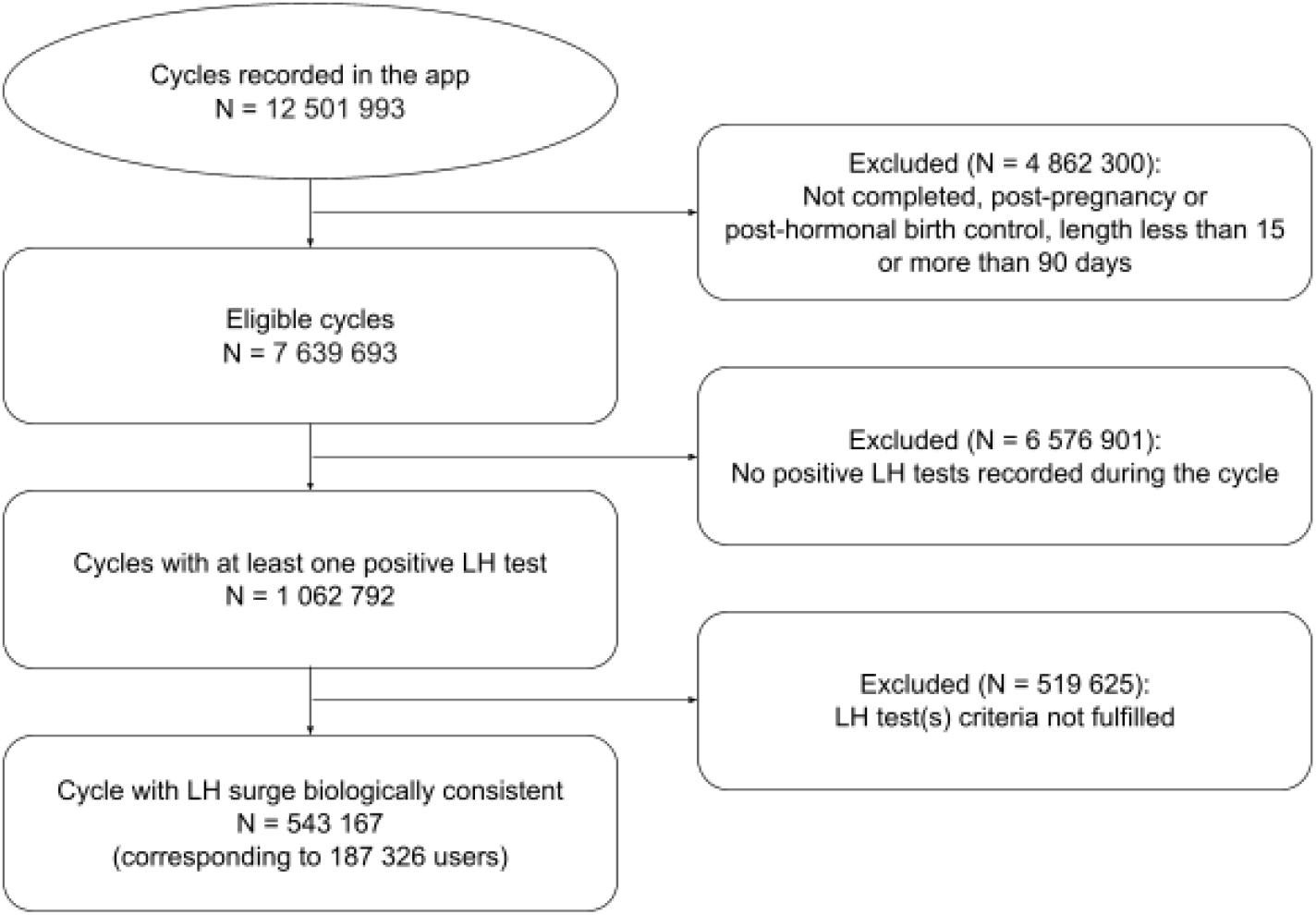
Flow diagram.

### Data collection and materials

The dataset included daily records of menstrual bleeding, basal body temperature measurements, and results of home urine luteinizing hormone (LH) tests. Basal body temperature (BBT) was measured using digital thermometers or wearable temperature-sensing devices supported by the application. Cycle-level variables comprised cycle start and end dates, and the user’s age at the beginning of each cycle. Additional user-level variables included country of residence, based on self-reported location at the time of analysis, as well as individual average cycle length and cycle-length variability at sign-up.

### Data analysis

The objective of the data analysis was to assess the agreement between days classified as fertile by typical period-tracking applications—considering both calendar-based approaches and approaches incorporating BBT—and the fertile days identified using urinary luteinizing hormone (LH) test results. The same evaluation was additionally performed for a digital birth control method based on BBT.

For all three methods, each cycle day was retrospectively classified as either unsafe (fertile) or safe (non-fertile) according to the method-specific fertile-window definition. Unsafe days correspond to days on which pregnancy is considered possible, while safe days correspond to days classified as having negligible pregnancy risk. We use the terms “safe” and “unsafe” here for consistency across methods and to reflect how users relying on these applications for pregnancy prevention may interpret these days, rather than the terminology used by individual applications.

All analyses were performed on the full set of eligible cycles and were additionally repeated in a predefined subgroup of irregular cycles, defined as cycles shorter than 21 days or longer than 35 days.

All approaches were implemented using the same underlying proprietary algorithm used in the Natural Cycles application to estimate the most likely ovulation day for each cycle. However, the input data provided to the algorithm varied depending on the method under consideration: calendar-based tracking used period dates only, whereas BBT-supported tracking and digital contraceptive used both period dates and basal body temperature data.

### Reference fertile window

To evaluate agreement between method-assigned fertile days (i.e. safe/unsafe days) and biological fertility, a reference fertile window representing the days on which conception is possible was defined.

The reference fertile window was based on LH testing. Ovulation was estimated to occur two days after the detected LH surge, consistent with evidence that ovulation typically follows the surge by approximately 24–48 hours [6]. The reference fertile window was defined as the six-day interval from day −5 to day 0 relative to this estimated ovulation day, reflecting evidence that the probability of conception outside this window is negligible. This LH-based fertile window served as the reference standard for evaluating agreement with fertile days assigned by each approach.

LH test information was used solely for the purpose of defining the reference fertile window and was not used as input to any of the fertile window estimation methods described below.

### Calendar-based period tracking

The calendar-based period-tracking approach estimates ovulation timing using menstrual cycle history, including cycle start dates and historical cycle lengths. Ovulation timing is estimated at the start of each cycle based solely on prior menstrual information.

Fertile status is assigned using a fixed seven-day fertile window, consistent with common practice in period-tracking applications. This window begins five days prior to the estimated ovulation day and ends one day after ovulation (days −5 to +1), with all days within this interval classified as unsafe.

### BBT-supported period tracking

The BBT-supported period-tracking approach extends the calendar-based method by incorporating historical BBT patterns from previous cycles to refine ovulation timing. No BBT-only approach (i.e. without menstrual dating) was evaluated, as commercially available BBT-supported period trackers typically also require period dates to enable predictions.

As with the calendar-based approach, fertile status is initiated five days prior to the estimated ovulation day. However, rather than using a fixed fertile window, fertile status is maintained until ovulation can be retrospectively confirmed by a sustained post-ovulatory temperature rise. All days classified as fertile during this interval are considered unsafe.

### FDA-cleared digital contraceptive

The FDA-cleared digital contraceptive approach applies the Natural Cycles algorithm using basal body temperature data and historical cycle information, without incorporating LH test results. For each cycle, days are classified by the algorithm as either unsafe (fertile) or safe (non-fertile) based on estimated ovulation timing and associated prediction uncertainty.

Unlike the period-tracking approaches, fertile status is not defined by a fixed window or a retrospective confirmation rule. Instead, safe and unsafe days are assigned dynamically on a day-by-day basis, accounting for uncertainty in ovulation estimation, which incorporates information on cycle regularity, data completeness, and factors known to affect menstrual cycle characteristics.

The sequence of days classified as unsafe within each cycle was taken to define the method-specific fertile window.

All data processing and analyses were performed using Python (version 3.10) and standard scientific computing libraries. Descriptive statistics were used to summarise user demographics and cycle characteristics.

### Outcome measures

An at-risk day was defined as a fertile day that was classified as safe by a given approach. High-risk days were defined as a subset of at-risk days corresponding to the two days with the highest probability of conception within the fertile window (days −2 and −1 with respect to ovulation).

Three outcome measures were evaluated:

- *Day-specific frequency of at-risk days*: For each day within the reference fertile window (days −5 to 0), we estimated the proportion of cycles in which that day was classified as safe by the method, thereby constituting an at-risk day.
- *Cycle-level misclassification of fertile days:* the percentage of cycles containing at least one at-risk day, as well as the percentage of cycles containing at least one or both high-risk days (as defined above).
- *Predicted pregnancy risk:* For each cycle, predicted pregnancy risk was calculated by summing, over all days in the reference fertile window, the probability of conception given intercourse for each day classified as at risk, using established day-specific probabilities of conception [10].

All outcome measures were additionally evaluated in a subgroup analysis restricted to irregular cycles, defined as cycles shorter than 21 days or longer than 35 days.

## Results

### Study population

After assessing menstrual cycles against the inclusion criteria, 543,167 cycles contributed by 187,326 users were included in the final analyses (Figure 2). A subset of included cycles were considered to be irregular (N = 26,325), the majority of which were longer than 35 days (N = 25,342), with a smaller proportion shorter than 21 days (N = 983).

The mean age of users at the start of a cycle was 30.0 years. The majority of users resided in the United States (58%), followed by the United Kingdom (19%) and Sweden (8%). At the user level, the mean average cycle length was 29.6 days, with a mean root mean square (RMS) cycle-length variability of 3.6 days.

### Day-specific frequency of at-risk days

Figure 3 shows the probability that a given cycle day was classified as unsafe relative to the LH-defined ovulation day for each method, together with the published day-specific probability of conception across the fertile window [10]. The calendar-based period tracker provided the fewest unsafe days, failing to cover the fertile window, and including days around ovulation.

**Figure 3.**
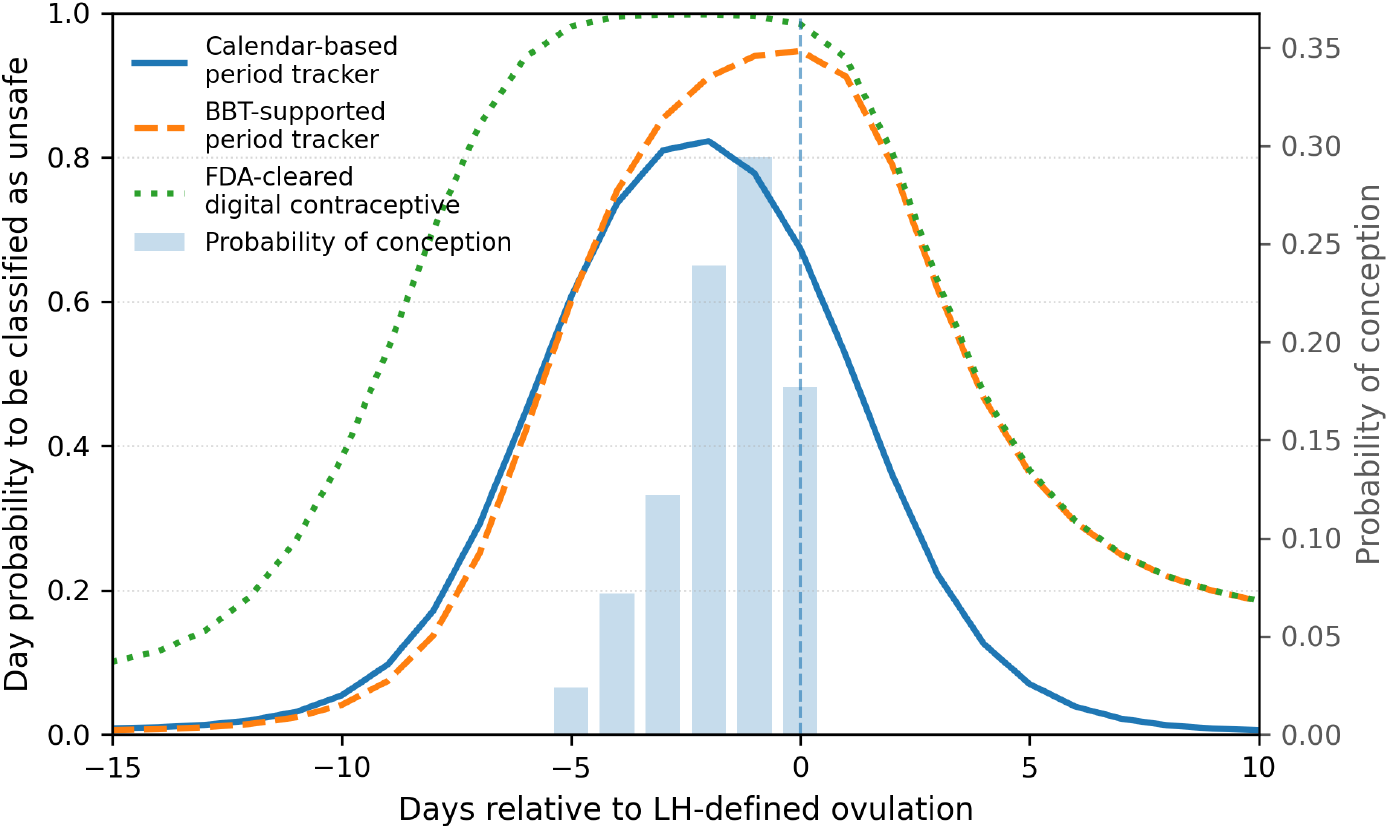
Probability that a given cycle day relative to the reference ovulation was classified as unsafe for each method, shown alongside the published day-specific probability of conception across the fertile window.

The BBT-supported period tracker closely resembled the calendar-based approach in the early pre-ovulatory phase, with days classified as unsafe at similar rates. However, unsafe classification remained elevated for longer around ovulation and, following ovulation, converged more closely with the FDA-cleared digital contraceptive.

The FDA-cleared digital contraceptive showed the most extensive coverage of days classified as unsafe, spanning the full interval of elevated conception probability shown in Figure 3. Overall, the timing and extent of days classified as unsafe differed across methods, with the digital contraception approach providing the most complete coverage of days associated with pregnancy risk.

To better visualise how often fertile days were misclassified as safe, Figure 4 shows the proportion of cycles with at-risk days as a function of the day relative to reference ovulation.

**Figure 4.**
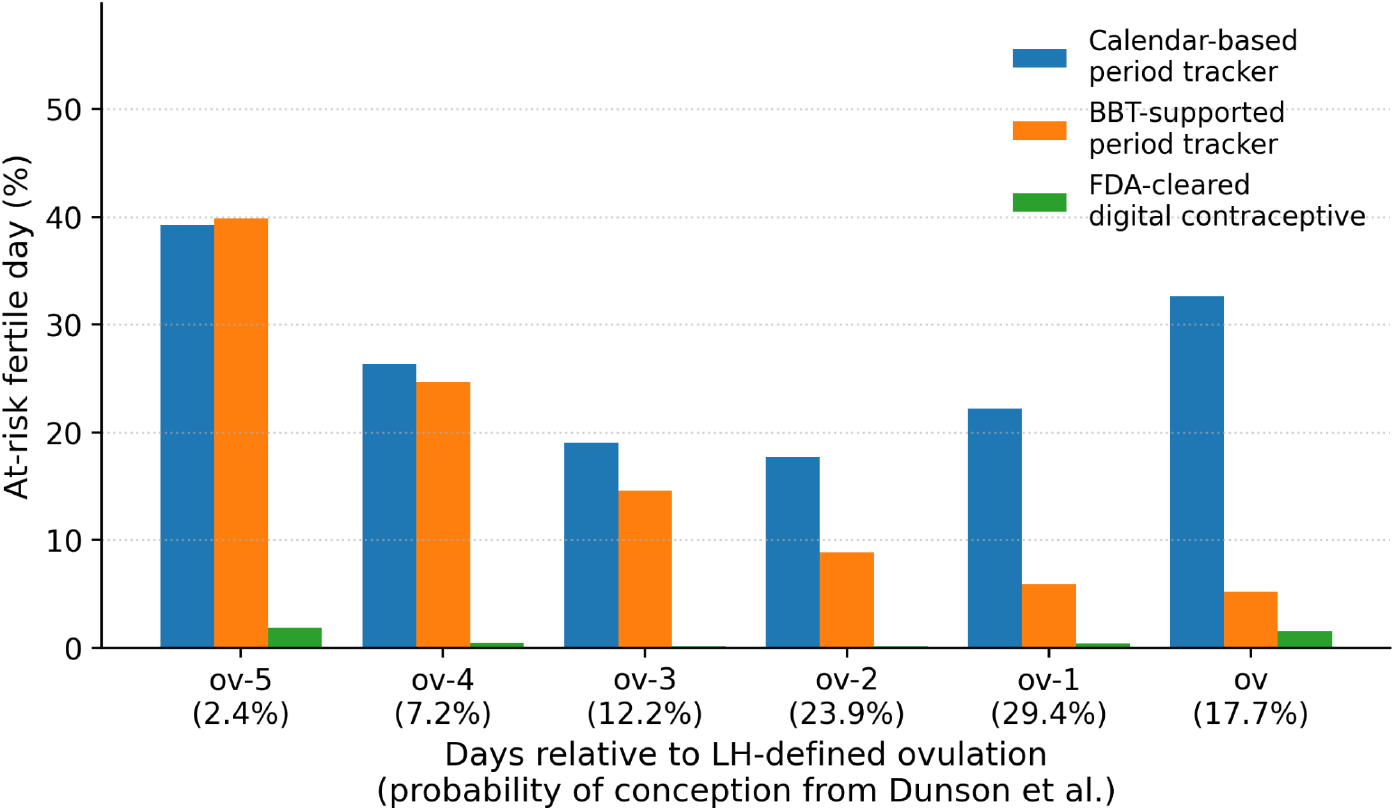
Proportion of cycles with fertile days misclassified as safe (at-risk days) as a function of the day relative to ovulation. The conception probability on each day is shown in brackets underneath.

The calendar-based period tracker showed consistently high frequencies of at-risk days across the fertile window, affecting approximately 40% of cycles at day −5 and remaining substantial on days of highest pregnancy risk (approximately 20%).

The BBT-supported period tracker showed lower frequencies of at-risk days on high-risk days compared with the calendar-based approach, with approximately 9% of cycles affected two days before ovulation (day −2) and 6% one day before ovulation (day −1).

The FDA-cleared digital contraceptive exhibited the lowest frequency of at-risk days across the fertile window, with misclassification remaining below 2% throughout.

### Cycle-level misclassification of fertile days

Table 1 summarises the proportion of cycles in which at least one at-risk day was observed, as well as the proportion of cycles in which one or two high-risk days were observed.

**Table 1.** Number and percentage of cycles with at least one at-risk day, at least one high-risk day and both high-risk days.

|  | Calendar-based period tracker | BBT-supported period tracker | FDA-cleared digital contraceptive |
| --- | --- | --- | --- |
| All cycles |  |  |  |
| Total number of cycles | 543,167 (100%) | 543,167 (100%) | 543,167 (100%) |
| Cycles with at least one at-risk fertile day | 365,330 (67%) | 224,046 (41%) | 18,107 (3%) |
| Cycles with at least one high-risk fertile day | 137,942 (25%) | 49,885 (9%) | 2,268 (0.4%) |
| Cycles with both high-risk fertile days | 78,790 (15%) | 31,070 (6%) | 668 (0.1%) |
| Irregular cycles |  |  |  |
| Total number of cycles | 26,325 (100%) | 26,325 (100%) | 26,325 (100%) |
| Cycles with at least one at-risk fertile day | 23,707 (90%) | 6,762 (26%) | 988 (4%) |
| Cycles with at least one high-risk fertile day | 19,963 (76%) | 3,388 (13%) | 293 (1%) |
| Cycles with both high-risk fertile days | 17,537 (67%) | 2,715 (10%) | 131 (0.5%) |

The calendar-based period tracker showed the highest frequency of misclassification. At least one at-risk day occurred in 67% of cycles, and 25% of cycles had at least one high-risk day classified as safe. Notably, both high-risk days were misclassified as safe in 15% of cycles, indicating that misclassification frequently affected the days with the highest probability of conception.

The BBT-supported period tracker showed lower rates of cycle-level misclassification. At least one at-risk day was observed in 41% of cycles, while at least one high-risk day was misclassified as safe in 9% of cycles. Misclassification of both high-risk days occurred in 6% of cycles.

The FDA-cleared digital contraceptive exhibited the lowest rates of misclassification. At least one at-risk day was observed in approximately 3% of cycles, and misclassification of at least one high-risk day occurred in less than 1% of cycles. Both high-risk days were misclassified as safe in 0.1% of cycles.

### Predicted pregnancy risk

To quantify potential pregnancy risk, we compared mean predicted pregnancy risk per cycle across methods, using day-specific probabilities of conception and assuming unprotected intercourse on at-risk days [10].

The calendar-based period tracker showed a mean predicted pregnancy risk of 22% per cycle, compared with 9% for the BBT-supported period tracker. The FDA-cleared digital contraceptive exhibited substantially lower predicted pregnancy risk, with a mean value of 0.5%.

### Subset analysis: irregular cycles

Patterns of fertile-day misclassification across methods were similar to those observed in the full study population, with higher absolute levels of misclassification. In irregular cycles, the calendar-based period tracker showed high rates of cycle-level misclassification. At least one at-risk day occurred in approximately 90% of cycles, and 76% of cycles had at least one high-risk day (day −2 or −1) classified as safe. Both high-risk days were misclassified as safe in 67% of irregular cycles.

The BBT-supported period tracker showed lower misclassification rates in this subgroup. At least one at-risk day occurred in 26% of cycles, and misclassification of at least one high-risk day was observed in 13% of cycles. The FDA-cleared digital contraceptive showed consistently low misclassification, with at-risk days present in 4% of cycles and misclassification of high-risk days occurring in ≤1% of cycles.

For irregular cycles, the mean predicted pregnancy risk per cycle for the calendar-based period tracker increased to 65%, whereas predicted pregnancy risk for the BBT-supported period tracker and the digital contraceptive were found to be at 12% and 1%, respectively.

## Discussion

### Findings and interpretation

In this large observational analysis, we quantified the extent to which fertile windows predicted by commonly used period-tracking apps fail to capture fertile days, as identified using LH testing. The analysis included 543,167 completed cycles with a well-defined LH surge pattern. Across all evaluated outcomes, both calendar-based and BBT-supported period-tracking apps were associated with frequent misclassification of fertile days as safe, including days with the highest probability of conception.

Under the calendar-based period tracker, 67% of cycles contained at least one at-risk day, and 25% contained at least one high-risk day (days −2 or −1) classified as safe. These misclassifications translated into a mean predicted pregnancy risk per cycle of approximately 22%.

Although incorporation of body temperature data reduced fertile-day misclassification relative to calendar-based tracking, substantial residual risk remained. BBT-supported period tracking improves post-ovulatory classification but lacks a mechanism to anticipate uncertainty before ovulation, and therefore still misclassifies pre-ovulatory fertile days as safe. Under the

BBT-supported period tracker, 41% of cycles still contained at least one at-risk day and 9% contained at least one high-risk day classified as safe, with a mean predicted pregnancy risk per cycle of approximately 9%. When such per-cycle pregnancy risk persists across multiple cycles, the cumulative likelihood of pregnancy increases substantially over time. These findings indicate that, while BBT-supported period tracking performs better than calendar-based tracking, it does not provide reliable protection against unintended pregnancy. Although period-tracking applications are not marketed or validated as contraceptive tools, the inclusion of features such as fertile-window predictions and ovulation estimates suggests that developers may be aware of—or even implicitly catering to—contraceptive use cases, without accepting the regulatory and validation responsibilities that this would entail.

The FDA-cleared digital contraceptive showed consistently low rates of fertile-day misclassification, with at least one at-risk day occurring in approximately 3% of cycles and high-risk day misclassification occurring in 0.4% of cycles. Predicted pregnancy risk per cycle was found to be 0.5%, indicating a meaningful difference between period-tracking apps and regulated digital contraceptives.

Irregular cycles further amplified the limitations of period-tracking applications. In this subgroup, calendar-based period tracking was associated with a marked increase in predicted pregnancy risk, reaching 65%. Differences for the BBT-supported period tracker and the digital contraceptive were less pronounced, likely because most irregular cycles in the dataset were long cycles, for which waiting for a sustained post-ovulatory rise in BBT helps reduce the risk of misclassification.

### Results in the Context of What is Known

To our knowledge, no prior studies have quantitatively assessed pregnancy risk arising from fertile-window misclassification in period-tracking applications using a biological ovulation reference. Existing literature has primarily focused on ovulation detection accuracy or on user perceptions and behaviours related to menstrual tracking [11]. Several qualitative and survey-based studies have reported that some women use period-tracking applications to inform contraceptive decisions, despite these tools not being designed or validated for pregnancy prevention [1,2].

Our findings extend this literature by providing a quantitative assessment of how fertile-window predictions from period-tracking apps translate into biologically meaningful pregnancy risk. Both calendar-based and BBT-supported period tracking applications frequently classified fertile days as safe, including days with high conception probability, thereby creating periods of unrecognised pregnancy risk. This mechanism offers a biologically grounded explanation for concerns raised in prior work regarding the use of period-tracking applications for pregnancy prevention.

In contrast, digital contraceptives based on BBT are regulated medical devices with documented contraceptive effectiveness and clearly defined use conditions. Our results show that this distinction has practical implications, translating into substantial differences in fertile-day coverage and predicted pregnancy risk when compared with period-tracking apps.

### Clinical Implications

These findings have direct implications for clinical counselling. Period-tracking applications are increasingly used in clinical practice, not only by patients independently but also on the recommendation of healthcare professionals for purposes such as cycle monitoring (variability and frequency) and symptom tracking (e.g. identifying patterns in conditions like PMDD). In doing so, clinicians may be passively endorsing these tools without providing explicit guidance on their contraceptive limitations.

This analysis clearly shows that period-tracking applications should not be considered reliable contraceptive tools. Although BBT-supported period tracking reduces risk relative to calendar-based tracking, it still leaves a substantial proportion of cycles with unrecognised exposure to pregnancy risk. This risk is even more pronounced for individuals with irregular cycles.

Healthcare professionals should be aware of the fundamental distinction between period-tracking applications and regulated digital contraceptives and should make clear to patients that any fertility-related predictions provided by the former should not be relied upon as medical advice for contraceptive decision-making. Digital contraceptives are designed for contraceptive use, taking uncertainty into account and widening the window of unsafe days when there are cycle irregularities or uncertainties in the data. These methods undergo regulatory review, publish effectiveness estimates, adjust their predictions over time as more user data becomes available, and communicate uncertainty and appropriate use conditions, such as including recommendations for abstinence or barrier method use. In contrast, most period-tracking applications—whether calendar-based or BBT-supported—are classified as wellness tools and are not required to demonstrate safety or effectiveness for pregnancy prevention. Clear differentiation during contraceptive counselling and outlining the risk of unintended pregnancy is essential, particularly for individuals seeking effective non-hormonal contraceptive options.

### Research Implications

These findings highlight the need for more rigorous evaluation and scrutiny of period tracking applications that present fertility-related predictions. Future research should move beyond assessing ovulation prediction accuracy alone and instead evaluate fertile-window misclassification and associated pregnancy risk.

Further work is needed to characterise how users interpret and act on fertile-window information provided by period-tracking applications, including how perceived “safe days” influence intercourse timing and contraceptive behaviour. Understanding these behavioural pathways is essential for quantifying the real-world impact of relying on period tracking applications for pregnancy prevention.

Finally, given the pronounced increase in pregnancy risk observed in irregular cycles, future studies should explicitly examine how cycle variability affects fertile-window prediction and pregnancy outcomes, and whether existing period tracking applications adequately communicate uncertainty to users with variable cycles.

### Strengths and Limitations

A key strength of this study is the large real-world dataset comprising over 500,000 menstrual cycles with urinary LH test data, enabling a biologically grounded estimation of ovulation timing at scale. The availability of LH data provided an independent reference for identifying fertile days, rather than relying on cycle-length assumptions alone. The sample size allowed detailed assessment of day- and cycle-level misclassification, and supported subgroup analyses in users with irregular cycles.

Several limitations should be acknowledged. A substantial proportion of cycles with LH testing were excluded due to incomplete or ambiguous LH surge patterns, reflecting real-world testing behaviour. While this reduced the number of eligible cycles, a strict definition of the LH surge was necessary to ensure a reliable biological reference, and the final sample remained large.

Ovulation timing was inferred from LH testing rather than confirmed by luteal serum progesterone and/or ultrasound, which is not feasible at this scale. Although we evaluated generic categories of period-tracking apps, the specific algorithms used by other commercial applications are unknown; accordingly, all fertile-window strategies were evaluated within a single analytical framework. While this enhances internal comparability, findings may not generalise to all period-tracking products.

## Conclusion

In this large real-world analysis, both calendar-based and basal body temperature–based period-tracking apps frequently misclassified biologically fertile days as safe, including days with the highest probability of conception. Although incorporation of BBT reduced misclassification relative to calendar-based tracking, substantial pregnancy risk remained across cycles, particularly among individuals with irregular cycles. In contrast, a regulated digital contraceptive based on BBT showed consistently low fertile-day misclassification and near-complete coverage of days associated with pregnancy risk.

These findings underscore that period-tracking applications, regardless of the data they incorporate, should not be considered reliable tools for pregnancy prevention. Clear differentiation between wellness-oriented period tracking applications and regulated digital contraceptives is essential in both clinical counselling and public health communication to reduce the risk of unintended pregnancy.

## Data Availability

All data produced in the present study are available upon reasonable request to the authors

